# Epidemiological characteristics of newly diagnosed Graves’ disease following the widespread outbreak of COVID-19 in Guangzhou, China

**DOI:** 10.64898/2025.12.28.25343112

**Authors:** Yongjie Lu, Ruting Zhong, Wen Shi, Die Zhou, Yuting Ran, Qiaoqiao Yang, Danqi Xu, Huijia Lin, Lei Yao, Shouwei Liao, Xiaodan Zhang, Ling Li, Wangen Li, Zhuoqing Hu

## Abstract

**Objectives:** There is currently insufficient evidence linking COVID-19 infection with Graves’ disease (GD). Following the complete lifting of COVID-19 restrictions on December 13, 2022, widespread infection in Guangzhou provides a basis for this study. This research aims to investigate the correlation between COVID-19 infection and GD onset, explore the epidemiological characteristics of newly diagnosed GD post-infection, and offer a scientific basis for treatment.

**Methods:** The study population included 494 GD outpatients treated in the Department of Endocrinology at the Second Affiliated Hospital of Guangzhou Medical University from January 1 to June 30 each year between 2021 and 2023. They were divided into two groups: 2023 (N=219) and 2021-2022 (N=275), based on the time node of widespread COVID-19 infection in 2023. The new diagnosis rate, general clinical characteristics, and serological test results of GD patients were analyzed before and after the outbreak of COVID-19.

**Results:** Compared with the 2021-2022 group, the new diagnosis rate of GD patients in 2023 showed a significant increase (12.8% vs. 8.4%, *P*<0.001). Furthermore, there was a significant decrease in pre-treatment thyrotropin receptor antibody levels (*P*=0.01), white blood cell count (*P*=0.02), and neutrophil proportion (*P*=0.04), while there was a significant increase in the proportion of patients with a family history (*P*=0.047). Follow-up until June 30 of that year revealed that the proportion of newly diagnosed GD patients developing hypothyroidism during treatment in 2023 significantly increased compared to the 2021-2022 group (*P*<0.001).

**Conclusions:** After widespread infection of COVID-19, the diagnosis rate of newly diagnosed GD increased, which may influence the epidemiological characteristics of related GD patients before initial treatment and during treatment.

## Introduction

Graves’ disease (GD) is an autoimmune thyroid disease (AITD) characterized by hyperthyroidism, diffuse enlargement of the thyroid gland, exophthalmos, and other symptoms. The global prevalence of hyperthyroidism is 0.2-1.3% [[1]], while the prevalence in China is 0.8%, with more than 80% attributed to GD. It is currently believed that GD is induced by intrinsic factors such as heredity, pregnancy, and gender, as well as extrinsic factors such as smoking and excessive iodine intake. Additionally, studies have shown a relationship between the occurrence and development of GD and viral infections [[2]-[3]]. A study following the outbreak of Coronavirus Disease 2019 (COVID-19) revealed that mortality rates were higher in patients with thyroid dysfunction [[4]]. For this reason, although respiratory symptoms are the most prominent clinical manifestations of COVID-19, the involvement of other systemic organs should not be overlooked [[5]]. COVID-19 infection can cause a wide variety of thyroid dysfunctions. Studies have shown that COVID-19 can lead to subacute thyroiditis (SAT) [[6]-[7]] and the manifestation of thyrotoxicosis. Furthermore, COVID-19 may be a potential risk factor for hypothyroidism [[8]-[9]], such as Hashimoto’s hypothyroidism or central hypothyroidism. To date, COVID-19-associated SAT has been frequently reported worldwide. Although many studies have well described the relationship between the novel coronavirus and AITD, including thyroid disease, the pathogenic mechanism has not been fully elucidated [[10]-[11]]. However, the relationship between COVID-19 and GD has mostly been reported as individual cases, and there is insufficient evidence supporting the association between the two.

During the outbreak of COVID-19 in mainland China, the country adhered to a dynamic zero-COVID policy for epidemic prevention. Later, since China announced the complete lifting of COVID-19 restrictions on December 13, 2022, the number of people reporting positive nucleic acid tests (NATs) for the novel coronavirus has continued to rise in all provinces, leading to widespread infection among the public. In our previous clinical practice, we observed an increase in newly diagnosed GD outpatients since 2023, and their clinical characteristics also showed some differences. To this end, in this study, we collected clinical data of newly diagnosed GD patients from January 1 to June 30 in 2021-2023, grouped and analyzed them, taking the year 2023 as the time node, to investigate the changes in the proportion of newly diagnosed GD patients, as well as the basic epidemiological characteristics of GD before and during treatment, following the widespread infection of COVID-19, with a view to providing scientific guidance for the treatment of GD.

## Methods

### Study Design and Subject Data

This study was designed as a retrospective cohort study, with subjects being outpatients newly diagnosed with GD from January 1 to June 30 of each year from 2021 to 2023 at the Second Affiliated Hospital of Guangzhou Medical University. Our study was initiated in July 2023, so the data for newly diagnosed GD patients in 2023 only covers the first half of the year. To ensure better comparability between the 2021-2022 and 2023 groups, we chose to analyze data from the first half of each year. Newly diagnosed GD patients were defined as those who met the diagnostic criteria for GD [[12]] and were diagnosed for the first time without any prior treatment. The demographic, clinical, and laboratory data of the subjects were obtained from the electronic medical record system for outpatients at the Second Affiliated Hospital of Guangzhou Medical University. The collected patient data served as baseline data, including basic information (gender, age, and date of visit), general conditions (including the degree of thyroid enlargement, presence of exophthalmos, complication type, drug use, and family history), and laboratory tests (including six items of thyroid function, blood routine, and liver function). To protect patient privacy, sensitive information such as patients’ names was not collected. Taking January 1, 2023, as the time node, all selected patients were followed up until June 30 of that year. The period from 2021 to 2022 was regarded as the period before the widespread infection of COVID-19, while the year 2023 was regarded as the period of widespread infection of COVID-19, in order to explore the differences in epidemiological characteristics of newly diagnosed GD patients before and after the widespread infection of COVID-19 in Guangzhou (see Fig.1 for the technical route). We conducted telephone follow-ups with newly diagnosed Graves’ disease patients to confirm the timing of their COVID-19 infection. Follow-up of COVID-19 infection events and their timing was conducted through patient clinic visits or telephone. In the follow-up, those with increased serum TSH and decreased FT_4_ in laboratory tests were diagnosed with hypothyroidism, while those with increased serum TSH and normal FT_4_ were diagnosed with subclinical hypothyroidism.

**Fig 1.**
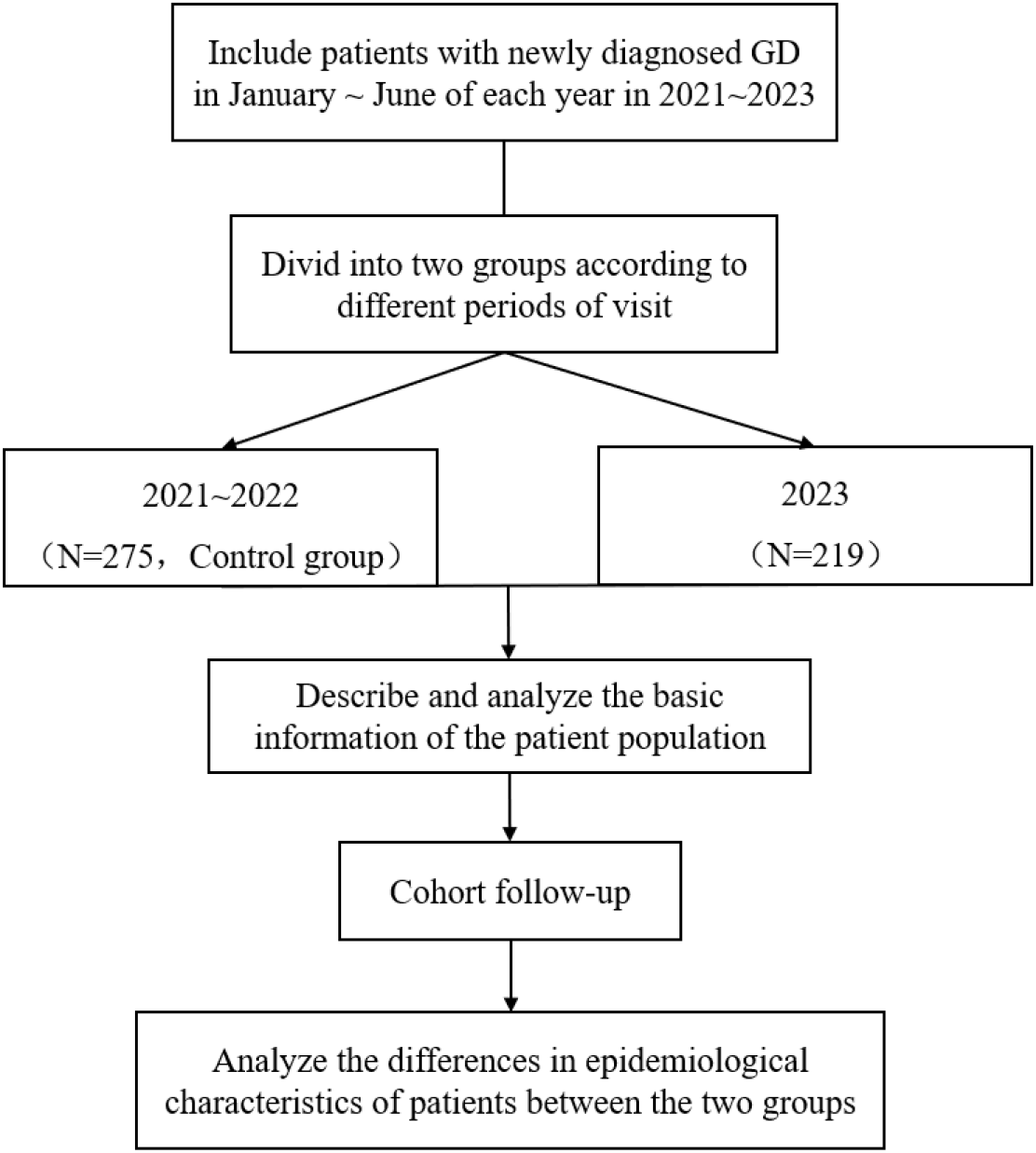
Flowchart of this Research

### Diagnosis of Graves’ disease

The diagnostic criteria for GD [[13]] are: ① Established diagnosis of hyperthyroidism; ② Diffuse goiter, with a minority of cases presenting without goiter; ③ Ophthalmopathy, such as eyelid retraction and other signs of Graves’ ophthalmopathy; ④ Skin and mucous membrane changes, such as pretibial myxedema or finger thickening; ⑤ Positive TRAb (third-generation method). Among the above criteria, items ① and ② are mandatory for the diagnosis, and the presence of at least one of items ③, ④, or ⑤ is sufficient for the diagnosis of GD. TRAb is the preferred serological test for the diagnosis of GD.

### Serological Test

All patients had their blood drawn for six items of thyroid function, blood routine, and liver function tests after fasting for 8-10 hours. Free triiodothyronine (FT_3_), free thyroxine (FT_4_), thyroid-stimulating hormone (TSH), thyroid peroxidase antibody (TPOAb), thyroglobulin antibody (TGAb), and thyrotropin receptor antibody (TRAb) were measured using the BCLIA method. Alanine aminotransferase (ALT) was measured using the rate method, and aspartate aminotransferase (AST) was measured using the colorimetric method. All of the above indicators were measured with a C8K instrument. White blood cell count (WBC), absolute neutrophil count (NEUT), and neutrophil ratio (NEUTr) were determined using an XN1000 instrument.

The normal ranges of serological test indicators were as follows: FT_3_ (3.1-6.8 pmol/L), FT_4_ (12-22 pmol/L), TSH (0.27-4.20 μIU/mL), TPOAb (0-34 IU/mL), TGAb (0∼115 IU/mL), TRAb (0-1.75 IU/L), ALT (5-40 U/L), AST (5-40 U/L), WBC (3.5-9.5 10^9^/L), NEUT (1.80-6.30 10^9^/L), NEUTr (40-75%).

### Statistical Analysis

We analyzed the pre-treatment test results of FT_3_, FT_4_, and TSH in newly diagnosed GD patients. TPOAb, TGAb, and TRAb were analyzed in two subgroups: those within the normal reference range and those greater than the normal range. For WBC, NEUT, and NEUTr, only differences in the proportion of patients below the normal range were explored. For ALT and AST, only differences in the proportion of patients exceeding the normal range were compared. For specific groupings of these indicators, please refer to Table 1.

**Table 1.**
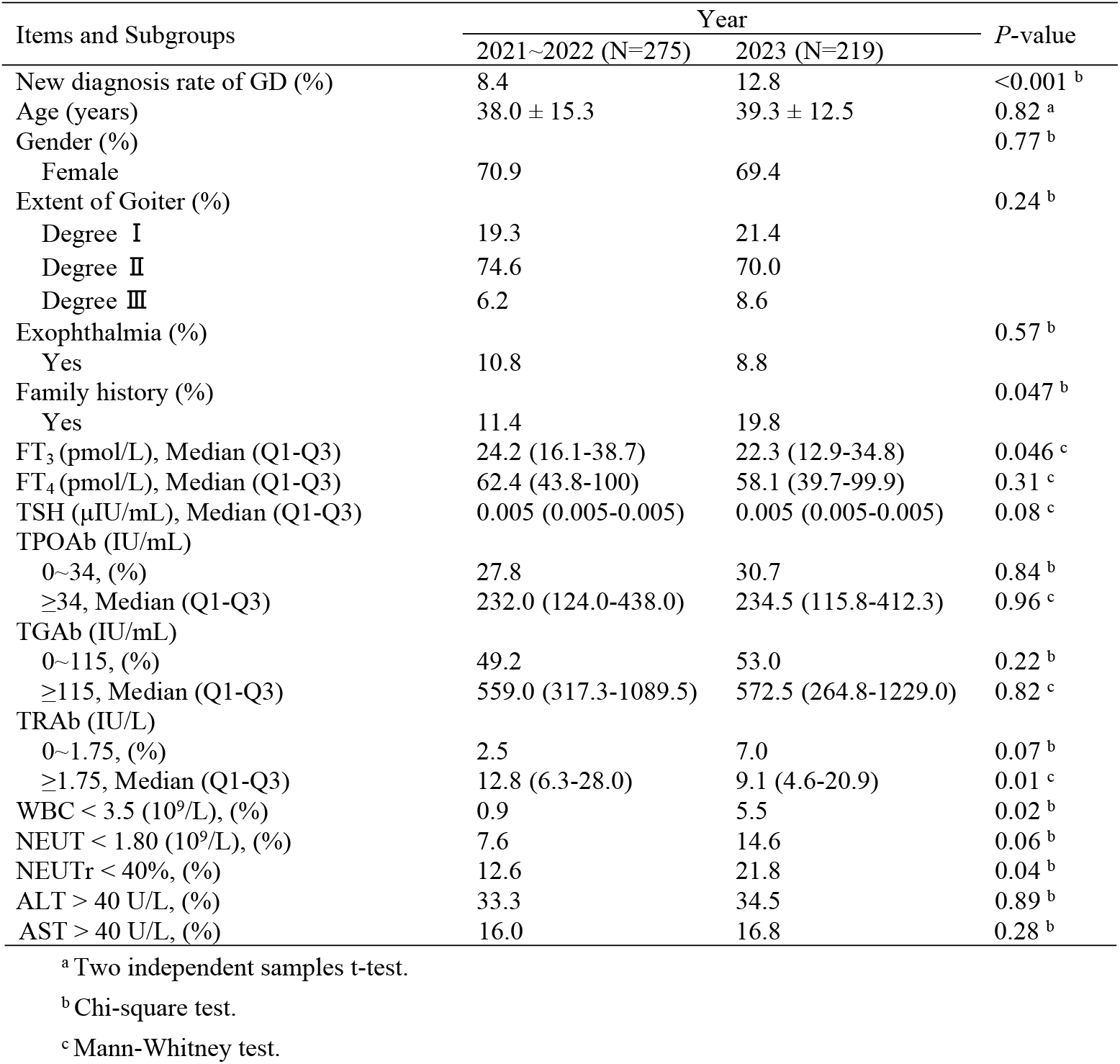
General Clinical Features of newly diagnosed GD patients before treatment.

| Items and Subgroups | Year |  | P-value |
| --- | --- | --- | --- |
|  | 2021~2022 (N=275) | 2023 (N=219) |  |
| New diagnosis rate of GD (%) | 8.4 | 12.8 | <0.001<sup>b</sup> |
| Age (years) | 38.0 ± 15.3 | 39.3 ± 12.5 | 0.82<sup>a</sup> |
| Gender (%) |  |  | 0.77<sup>b</sup> |
| Female | 70.9 | 69.4 |  |
| Extent of Goiter (%) |  |  | 0.24<sup>b</sup> |
| Degree I | 19.3 | 21.4 |  |
| Degree II | 74.6 | 70.0 |  |
| Degree III | 6.2 | 8.6 |  |
| Exophthalmia (%) |  |  | 0.57<sup>b</sup> |
| Yes | 10.8 | 8.8 |  |
| Family history (%) |  |  | 0.047<sup>b</sup> |
| Yes | 11.4 | 19.8 |  |
| FT<sub>3</sub> (pmol/L), Median (Q1-Q3) | 24.2 (16.1-38.7) | 22.3 (12.9-34.8) | 0.046<sup>c</sup> |
| FT<sub>4</sub> (pmol/L), Median (Q1-Q3) | 62.4 (43.8-100) | 58.1 (39.7-99.9) | 0.31<sup>c</sup> |
| TSH (μIU/mL), Median (Q1-Q3) | 0.005 (0.005-0.005) | 0.005 (0.005-0.005) | 0.08<sup>c</sup> |
| TPOAb (IU/mL) |  |  |  |
| 0~34, (%) | 27.8 | 30.7 | 0.84<sup>b</sup> |
| ≥34, Median (Q1-Q3) | 232.0 (124.0-438.0) | 234.5 (115.8-412.3) | 0.96<sup>c</sup> |
| TGAb (IU/mL) |  |  |  |
| 0~115, (%) | 49.2 | 53.0 | 0.22<sup>b</sup> |
| ≥115, Median (Q1-Q3) | 559.0 (317.3-1089.5) | 572.5 (264.8-1229.0) | 0.82<sup>c</sup> |
| TRAb (IU/L) |  |  |  |
| 0~1.75, (%) | 2.5 | 7.0 | 0.07<sup>b</sup> |
| ≥1.75, Median (Q1-Q3) | 12.8 (6.3-28.0) | 9.1 (4.6-20.9) | 0.01<sup>c</sup> |
| WBC < 3.5 (10<sup>9</sup>/L), (%) | 0.9 | 5.5 | 0.02<sup>b</sup> |
| NEUT < 1.80 (10<sup>9</sup>/L), (%) | 7.6 | 14.6 | 0.06<sup>b</sup> |
| NEUTr < 40%, (%) | 12.6 | 21.8 | 0.04<sup>b</sup> |
| ALT > 40 U/L, (%) | 33.3 | 34.5 | 0.89<sup>b</sup> |
| AST > 40 U/L, (%) | 16.0 | 16.8 | 0.28<sup>b</sup> |

<sup>a</sup> Two independent samples t-test.
<sup>b</sup> Chi-square test.
<sup>c</sup> Mann-Whitney test.

Normally distributed continuous variables were presented as mean ± SD, non-normally distributed continuous variables were presented as median (Q1-Q3), while categorical variables were presented as count (percentage). Differences in categorical variables were analyzed using the chi-square test, and normally distributed continuous variables were analyzed using the t-test. The Mann-Whitney test was used for non-normally distributed continuous variables. All tests were two-tailed, with P<0.05 indicating statistical significance. Empower (R) (www.empowerstats.com, X&Y Solutions, Inc., Boston, MA) and R (http://www.r-project.org) were used for all statistical analyses in this study, and GraphPad Prism 8.0.1 was adopted for plotting.

### Ethical approval

This study was approved by the Ethics Committee of the Second Affiliated Hospital of Guangzhou Medical University, China (2023-hs-11-02) on March 1, 2023. Written informed consent was obtained from all participants.

## Results

### General clinical characteristics of newly diagnosed GD patients before treatment

The total number of patients seeking medical attention for GD from 2021 to 2022 was 3,267, including 275 newly diagnosed GD patients. In 2023, the total number of GD patients was 1,707, including 219 newly diagnosed patients. The rate of loss to follow-up was less than 10% at the end of observation. According to the information provided by the patients, two participants in the 2023 cohort were clearly free of COVID-19, whereas no participants in the 2021-2022 cohort were infected with COVID-19 prior to enrollment. Compared with the 2021-2022 group, the diagnosis rate of newly diagnosed GD patients significantly increased in 2023 (12.8% vs. 8.4%, *P*<0.001, see Fig.2). The proportion of patients with a family history also significantly increased (19.8% vs. 11.5%, *P*=0.047). There were no differences between the two groups in terms of age, gender, degree of thyroid enlargement, and presence of exophthalmos. Compared with 2021-2022, the level of FT3 among newly diagnosed GD patients significantly decreased in 2023 (*P*=0.046), and FT_4_ showed a downward trend; however, the differences were not statistically significant(*P*=0.31).

**Fig 2.**
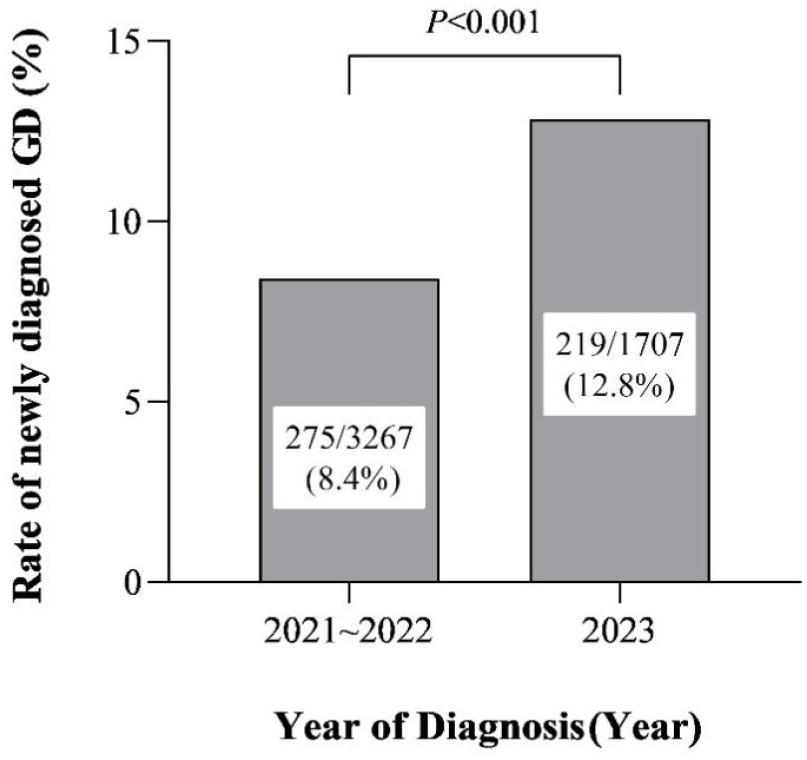
The diagnosis rate of newly diagnosed GD in 2021∼2023

Compared with the 2021-2022 group, the TRAb value in newly diagnosed GD patients in 2023 significantly decreased (*P*=0.01), and the proportion in the 0-1.75 IU/L subgroup (TRAb normal group) showed an upward trend, although it was statistically insignificant (*P*=0.07). The trends of TPOAb and TGAb in the two groups were consistent with that of TRAb; both were statistically insignificant.

An analysis of the blood routine and liver function test results indicated that the proportions of newly diagnosed GD patients whose WBC (*P*=0.02) and NEUTr (*P*=0.04) were below the normal range in 2023 were significantly higher than those from 2021-2022. There were no differences in ALT and AST (see Table 1 for details).

### Follow-up of newly diagnosed GD patients for hypothyroidism during treatment

In the subsequent follow-up, we found that the proportion of newly diagnosed GD patients who developed hypothyroidism within six months after diagnosis significantly increased in 2023 (17.4% vs. 5.8%, *P*<0.001), (see Table 2 and Fig.3).

**Table 2.** Follow-up of newly diagnosed GD patients for hypothyroidism during treatment, (%)

| | Year | | $\chi^2$-value | P-value |
| --- | --- | --- | --- | --- |
|  | 2021~2022 | 2023 |  |  |
| Non- hypothyroidism | 252 (91.6) | 178 (81.28) | 11.60 | <0.001 |
| Subclinical Hypothyroidism | 7 (2.6) | 3 (1.4) | 0.85 | 0.36 |
| Hypothyroidism | 16 (5.8) | 38 (17.4) | 16.66 | <0.001 |

**Fig 3.**
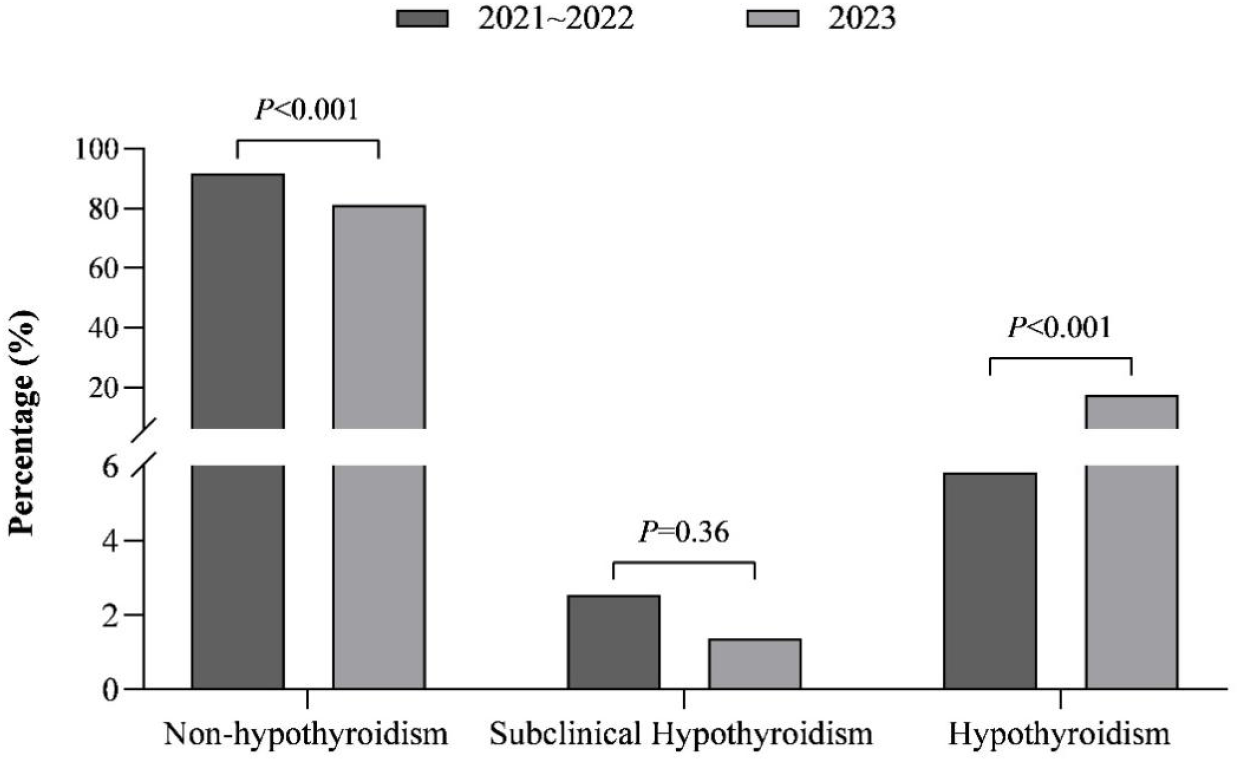
Follow-up of newly diagnosed GD patients for hypothyroidism during treatment

## Discussion

Our study found that following the complete lifting of COVID-19 restrictions and widespread infection of COVID-19 in the general public in China, there has been a significant increase in the diagnosis rate of newly diagnosed GD patients (12.8% vs. 8.4%), which may be closely related to COVID-19 infection. As demonstrated by studies, COVID-19 infection can induce a variety of autoimmune diseases, including AITD, and even result in GD [[14]-[15]]. Furthermore, two recent retrospective cohort studies this year also demonstrated a rise in the incidence of newly diagnosed GD in minors during the outbreak of COVID-19 and an increased risk of multiple autoimmune diseases, including GD [[16]-[17]]. Viral infection is considered one of the pathogenic mechanisms of AITD, and the pathological mechanism whereby COVID-19 infection triggers GD remains elusive. This is probably because the thyroid highly expresses angiotensin-converting enzyme 2 (ACE2) protein, through which COVID-19 directly enters the cells, causing direct damage to the thyroid [[18]-[21]], or because COVID-19 infection induces an increase in the levels of cytokines like IL-6, resulting in abnormal autoimmunity of the thyroid and indirectly leading to the occurrence of GD [[22]-[23]]. Secondly, COVID-19-associated GD can also be induced by the stress response and fear accompanying the diagnosis of COVID-19 in the early stage of the epidemic.

Through the analysis of the results of six items of thyroid function of newly diagnosed GD patients in this study, we found that the level of FT_3_ significantly decreased in 2023. Although there were no statistical significances in terms of FT_4_, FT_4_ also showed a downward trend. We also observed that the value of TRAb was significantly lower in 2023, and the proportion of TRAb-negative patients showed an upward trend, which may be one of the reasons for the downward trend of FT_3_ and FT_4_ in newly diagnosed GD patients in 2023 and be attributable to the delayed appearance of the TRAb antibody in COVID-19-induced GD patients. A case report described a female patient with newly diagnosed GD who tested positive for TRAb and TSI until 3 months after being infected with COVID-19. This is the longest delayed TRAb-positive case so far [[24]], highlighting the necessity to monitor the delayed positivity of the GD antibody. On the other hand, through follow-up, we discovered that the proportion of newly diagnosed GD patients who developed hypothyroidism within six months after diagnosis significantly increased in 2023 (18.7% vs. 5.8%), which may be closely related to the lower TRAb level in newly diagnosed GD patients. TRAb is the most specific and best-correlated antibody for the diagnosis of GD, with more than 95% of newly diagnosed patients testing positive. TRAb positivity is regarded as the pathogenic antibody of GD, and its level also partly influences the level of thyroid hormones. It is an important indicator to judge the risk of relapses and drug discontinuance during treatment. The subjects of this study were treated with either methimazole or propylthiouracil. Since our medication was initiated and adjusted according to the guideline, the possibility of drug-induced hypothyroidism was low. There were isolated reports that after the initiation of methimazole treatment in GD patients following COVID-19 infection, symptoms relieved quickly and reached the indication for discontinuation [[25]], which coincides with our observation of short-term occurrence of hypothyroidism in newly diagnosed GD patients after COVID-19 infection, but the specific mechanism is yet to be confirmed. In clinical practice, if a patient has a clear history of COVID-19 before diagnosis, physicians should remain vigilant for the emergence of hypothyroidism during treatment and adjust doses in time accordingly.

It has been reported that COVID-19 infection mostly results in an increase in the level of inflammatory cytokines and WBC [[26]-[27]]. Leukopenia or aleukemia and neutropenia are common complications of GD, occurring before treatment and after antithyroid drug treatment [[28]-[29]]. For this reason, the phenomenon of more patients with WBC, NEUT, and NEUTr below the normal range in newly diagnosed GD patients in 2023 may be strongly related to GD, possibly attributed to the combined effects of autoimmune reactions directly caused by COVID-19 and excessive thyroid hormone on the hematopoietic system.

Family clustering is one of the features of GD. It is generally held that GD is a complex disease with multiple pathogenic factors. The results of twin cohort studies show that about 80% of GD may be attributed to genetic factors, while viral infection and other environmental factors account for only about 20% [[30]]. However, the currently identified susceptibility genes for GD have limited explanatory power and struggle to account for familial clustering. The study of gene-environment interaction may be an important approach to enhance our understanding of the pathogenesis of GD. Our study found that the proportion of newly diagnosed patients with a family history of GD significantly grew in 2023 (19.8% vs. 11.5%). Apart from COVID-19 infection, interaction with a positive family history of GD may also be an important reason for the high diagnosis rate of newly diagnosed GD in 2023. However, the present study had a relatively short follow-up period and didn’t gather specific information regarding positive family history. Further evidence from follow-up and more studies is needed to confirm the role of the interaction between COVID-19 infection and genetic factors in the pathogenesis of GD, and explore its role in the progression of the disease.

Taken together, the global outbreak of COVID-19 has partly induced the occurrence and development of thyroid diseases. Our study suggested that after the widespread infection of COVID-19 in China, the diagnosis rate of newly diagnosed GD increased, indicating that COVID-19 infection may be a risk factor for GD. It presents related epidemiological characteristics and provides reference for the clinical treatment of GD. There are some limitations to this study: it was carried out in a single center, and the results may not be extrapolated in a full and accurate manner. However, our hospital is a large Grade III Level A hospital in Guangzhou, and our findings may reflect the real activity of the disease in this region. This study only analyzed baseline data of two different period cohorts. To explore the epidemiological characteristics during the treatment and after drug withdrawal, further follow-up is needed to acquire the subsequent general condition and laboratory test results of the patients. Hence, more multicenter studies are needed to clarify the correlation between COVID-19 infection and GD. Lastly, this study did not collect information on COVID-19 vaccination for each patient, thus we were unable to analyze the potential association between vaccination status and Graves’ disease (GD). Although COVID-19 vaccination was free and widely administered in China under national policy, excluding special groups such as pregnant women and children, the vaccination rate may have been similar between the 2021-2022 group and the 2023 group. Future research will focus on the impact of COVID-19 vaccination on GD.

## 5. Conclusion

After the widespread infection of COVID-19, the diagnosis rate of newly diagnosed GD increased, which may influence the epidemiological characteristics of related GD patients before and during treatment.

## Ethical statement

This study has been approved by the Ethics Committee of the Second Affiliated Hospital of Guangzhou Medical University, China (2023-hs-11-02).

## Data availability statement

The datasets supporting the findings of current study are available from the corresponding author on reasonable request.

## Author contributions

**Y.J. Lu and R.T. Zhong** conducted literature reviews, participated in data analysis, contributed to data interpretation, and wrote the first draft of the manuscript. **D. Zhou and X.D. Zhang** participated in patient registration, enrollment and data collection. **W. Shi, Y.T. Ran, L. Yao and S.W. Liao** participated in the retrieval and entry of patient data. **Q.Q. Yang, D.Q. Xu and H.J. Lin** participated in the data analysis and contributed to the data interpretation. **L. Li and W.G. Li** contributed to the interpretation of the data and critically reviewed the manuscript. **Z.Q. Hu** contributed to the conception and design of this study, contributing to the specific critical review of the manuscript. All authors revised, read, and approved the final manuscript.

## Acknowledgements

The authors thank all patients for participating in this study. Data from the Second Affiliated

Hospital of Guangzhou Medical University was obtained.

